# Prioritization of risk genes for Alzheimer’s disease: an analysis framework using spatial and temporal gene expression data in the human brain based on support vector machine

**DOI:** 10.1101/2023.02.06.23285522

**Authors:** Shiyu Wang, Xixian Fang, Xiang Wen, Congying Yang, Ying Yang, Tianxiao Zhang

**Affiliations:** Department of Epidemiology and Biostatistics, School of Public Health, Xi’an Jiaotong University Health Science Center; Hangzhou Institute for Advanced Study, University of Chinese Academy of Sciences, Beijing, China; National Anti-Drug Laboratory Shaanxi Regional Center, Xi’an, China

**Author notes:** ***Correspondence:*** Tianxiao Zhang, Tang Scholar, Department of Epidemiology and Biostatistics, School of Public Health, Xi’an, Jiaotong University Health Science Center, Xi’an, Shaanxi, 710061, China.

**Keywords:** Alzheimer’s disease, risk gene prioritization, gene expression patterns, machine learning, genome-wide association analyses

## Abstract

**Background:** Alzheimer’s disease (AD) is a complex disorder, and its risk is influenced by multiple genetic and environmental factors. In this study, an AD risk gene prediction framework based on spatial and temporal features of gene expression data (STGE) was proposed.

**Methods:** We proposed an AD risk gene prediction framework based on spatial and temporal features of gene expression data. The gene expression data of providers of different tissues and ages were used as model features. Human genes were classified as AD risk or non-risk sets based on information extracted from relevant databases. Support vector machine (SVM) models were constructed to capture the expression patterns of genes believed to contribute to the risk of AD.

**Results:** The recursive feature elimination (RFE) method was utilized for feature selection. Data for 64 tissue-age features were obtained before feature selection, and this number was reduced to 19 after RFE was performed. The SVM models were built and evaluated using 19 selected and full features. The area under curve (AUC) values for the SVM model based on 19 selected features (0.740 [0.690–0.790]) and full feature sets (0.730 [0.678–0.769]) were very similar. Fifteen genes predicted to be risk genes for AD with a probability greater than 90% were obtained.

**Conclusion:** The newly proposed framework performed comparably to previous prediction methods based on protein-protein interaction (PPI) network properties. A list of 15 candidate genes for AD risk was also generated to provide data support for further studies on the genetic etiology of AD.

## 1 Introduction

Alzheimer’s disease (AD) is a chronic neurodegenerative disorder that is characterized by cognitive impairment and memory loss. It affected approximately 50 million people worldwide in 2020, which is expected to increase to 150 million by 2050 [1]. Advanced age is the most important risk factor for AD [2]. A significant increase in the incidence rate of AD was observed in senior citizens after the age of 65 years [2]. Equal incidence rates of AD were identified for males and females after adjusting for age, indicating that sex might not be associated with the risk of AD [2]. The pathological features of AD include senile plaques formed by the accumulation of β-amyloid protein and neurofibrillary tangles composed of highly phosphorylated τ proteins. Several hypotheses have been proposed to explain the pathogenesis of AD, including oxidative stress [3], inflammation [3], and DNA damage [4]. However, no consensus has yet been reached.

Previous studies have indicated that AD is a complex disorder, and its risk is attributed to multiple genetic and environmental factors [5-6]. In the last decade, genome-wide association (GWA) analyses have significantly contributed to the genetic etiology of AD [6]. Jansen *et al*. confirmed 29 risk loci and several relevant pathways related to AD through a GWA meta-analysis [7]. In addition, Celeste *et al*. reviewed the relationship between several AD risk genes, including *ABCA7, BIN1, CASS4*, and *CD33*, and the cellular and neuropathological characteristics of AD [8]. Nevertheless, a recent study indicated that approximately half of the heritability of AD remains unaccounted [9]. It is probable that a large number of susceptibility loci for AD have not yet been discovered. However, recent studies have indicated that larger-scale GWA studies in the future are less cost effective due to the intrinsic deficiency rooted in the study design of GWA studies; therefore, it might not be a preferable choice for unraveling these hidden genomic regions that contribute to the risk of AD [10]. In this sense, prioritizing AD risk genes based on evidence gained from different perspectives and then validating these candidate risk genes in subsequent candidate gene-based association studies might be an effective strategy for discovering more relevant genes for AD risk. In a recent study, Cogill *et al*. applied machine-learning-based methods using brain developmental gene expression data to prioritize high-confidence candidate genes for autism spectrum disorder [11]. This study established a feasible analysis pipeline for prioritizing candidate risk genes for complex disorders, using spatial and temporal gene expression data.

Multiple lines of evidence have indicated that the expression of AD risk genes has specific spatial and temporal features [12-13]. Extracting and properly synthesizing information from these gene expression features might be an effective way to prioritize the risk genes for AD. In this study, we aimed to construct and evaluate a machine-learning-based model to identify high-confidence risk genes for AD using spatial and temporal gene expression data extracted from a publicly available database.

## 2 Methods

The statistical analysis pipeline is shown in Figure 1. In this study, we propose an AD risk gene prediction framework based on spatial and temporal features of gene expression data (STGE). In this analysis framework, the gene expression data of providers of different tissues and ages were utilized as model features. Human genes were classified as AD risk or non-risk sets and randomly split into training and validation sets. Support vector machine (SVM) models were constructed to capture the expression patterns of genes that were believed to contribute to the risk of AD in the training set, which were then applied to the validation set to evaluate model performance. The STGE model was then applied to a gene set with an unknown status for AD risk, and a confidence score was assigned to each gene.

**Figure 1.**
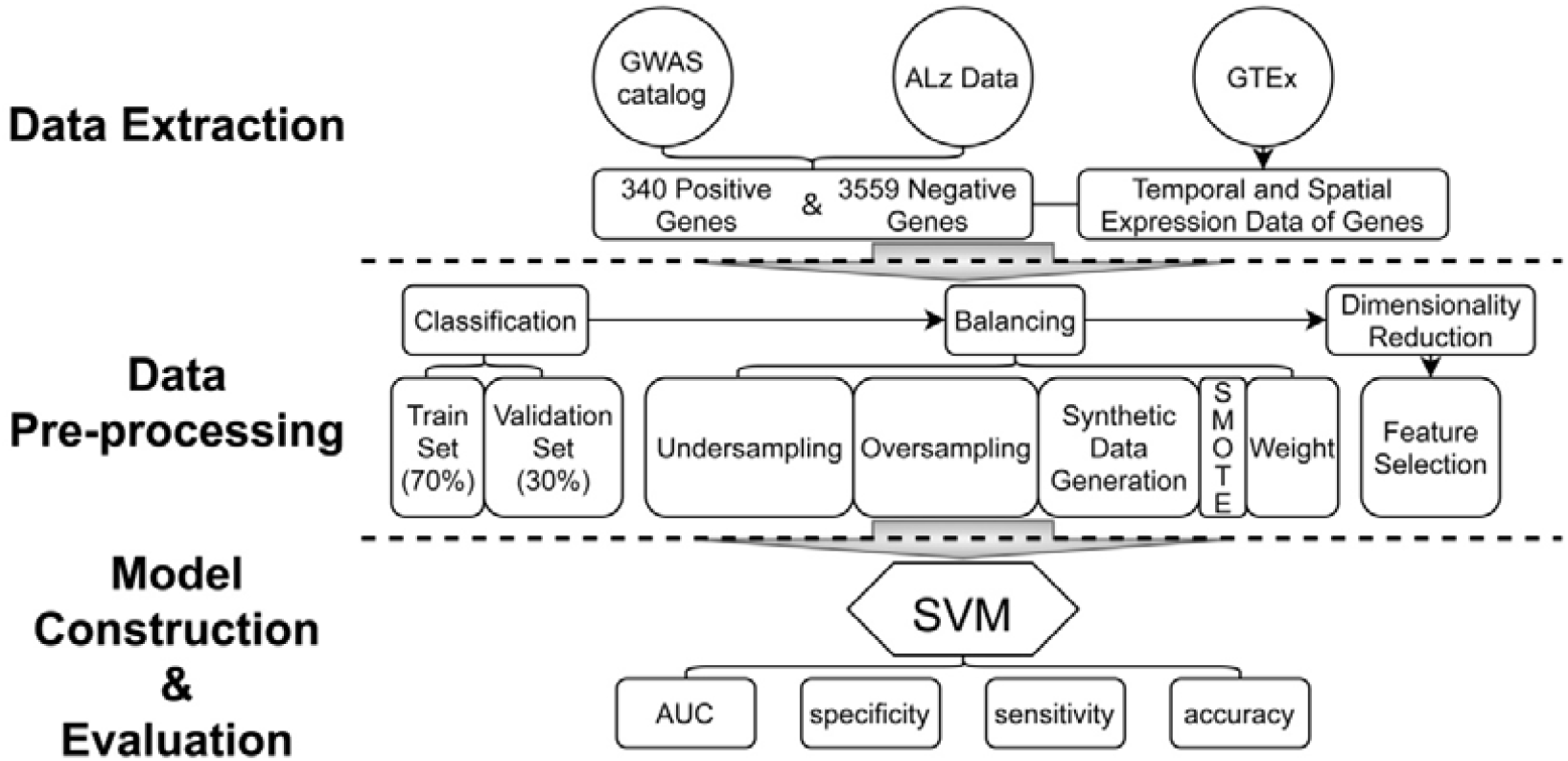
Analysis pipeline of the model construction and evaluations

### 2.1 Data Extraction

The data used in the present study were extracted from three publicly available databases: the GTEx database (https://gtexportal.org/home/) [14], AlzData database (http://www.alzdata.org/) [15], and GWAS catalog (https://www.ebi.ac.uk/gwas/) [16].

Spatial and temporal expression data for each gene were obtained from the GTEx database. Gene expression data related to tissues of the human brain (including the cerebellum, cortex, anterior cingulate cortex, hippocampus, substantia nigra, caudate, cerebellar hemisphere, frontal cortex, hypothalamus, nucleus accumbens, putamen, spinal cord, and amygdala) were extracted. Data from tissue sample providers under 20 or over 70 years of age were not included. In addition, we also removed tissue providers who scored 0 or 4 points on the 4-point Hardy Scale for their death classification. Finally, gene expression data in 13 types of brain-related tissues for 14,697 genes were extracted from 317 tissue sample providers of various ages and genders (Supplementary Table S1 and Supplementary Figure S1).

AlzData is a database for scoring correlations between human genes and the risk of AD, based on evidence from high-throughput omics data. The scores ranged from 0 to 5, with a higher score indicating a stronger correlation between the gene and AD. Genes with scores of 4 to 5 were extracted to form the AD risk gene set (“the right answer”). For genes with scores of 0 to 3, we supplemented the information from the GWAS catalog and excluded genes related to AD to obtain AD non-risk genes. Finally, 3,899 genes comprising 340 AD risk genes and 3,559 non-AD risk genes were identified.

### 2.2 Model Construction and Evaluation

The SVM models were constructed based on spatial and temporal gene expression data extracted from relevant databases using the e1071 package of the R project. Gene expression data were first grouped by the tissue type and age of the tissue providers. The median expression level of each gene in the tissue type age group was calculated and used as features in the SVM models. A total of 64 brain tissue-related features were obtained for model construction (Supplementary Table S2). The dataset was randomly divided into training and validation sets in a ratio of 7:3. There were 238 AD risk genes and 2,491 AD non-risk genes in the training set. The SMOTE function in the DMwR package was used to balance gene numbers. Feature selection was conducted using the caret package, and 19 features were selected based on recursive feature elimination (RFE). Accuracy and Kappa statistics were chosen as the evaluation indicators to estimate the performance of the selected features, and we chose the feature set with both the greatest value and least variance to build the SVM model. Parameter optimization was performed using a grid search strategy. Parameters including model accuracy, specificity, sensitivity, and area under the curve (AUC) were utilized to evaluate the performance of the SVM model. The R packages pROC and ROCR were used to draw the ROC curve and calculate the AUC, respectively. The R package ggplot2 was used for data visualization.

## 3 Results

### 3.1 Feature selection based on recursive feature elimination

RFE was used for feature selection. Data for 64 tissue-age features were obtained before feature selection, and this number was reduced to 19 after RFE was performed. SVM models based on each of these 19 features (the gene expression levels were obtained by median values of samples) were built and evaluated for accuracy, specificity, sensitivity, and AUC (Table 1). The feature with the highest AUC was the human tissue of the brain cerebellum at the age of 40–49 (AUC=0.688).

**Table 1.** The mean accuracy, sensitivity, specificity and AUC of each model built by each selected feature from the RFE method

| Tissue | Age | Accuracy | Sensitivity | Specificity | AUC |
| --- | --- | --- | --- | --- | --- |
| Brain-Cerebellum | 40-49 | 0.604 | 0.657 | 0.599 | 0.688 |
| Brain-Amygdala | 50-59 | 0.768 | 0.441 | 0.799 | 0.684 |
| Brain-Frontal Cortex BA9 | 50-59 | 0.657 | 0.598 | 0.663 | 0.683 |
| Brain-Anterior cingulate cortex BA24 | 30-39 | 0.67 | 0.559 | 0.681 | 0.683 |
| Brain-Putamen basal ganglia | 30-39 | 0.645 | 0.569 | 0.653 | 0.682 |
| Brain-Anterior cingulate cortex BA24 | 40-49 | 0.701 | 0.529 | 0.717 | 0.678 |
| Brain-Cerebellum | 60-69 | 0.551 | 0.676 | 0.539 | 0.674 |
| Brain-Cerebellar Hemisphere | 60-69 | 0.689 | 0.549 | 0.702 | 0.667 |
| Brain-Frontal Cortex BA9 | 60-69 | 0.644 | 0.520 | 0.656 | 0.665 |
| Brain-Substantia nigra | 40-49 | 0.715 | 0.500 | 0.736 | 0.664 |
| Brain-Putamen basal ganglia | 60-69 | 0.691 | 0.520 | 0.707 | 0.655 |
| Brain-Caudate basal ganglia | 30-39 | 0.695 | 0.559 | 0.708 | 0.640 |
| Brain-Anterior cingulate cortex BA24 | 50-59 | 0.733 | 0.461 | 0.759 | 0.636 |
| Brain-Cerebellum | 30-39 | 0.770 | 0.461 | 0.800 | 0.633 |
| Brain-Substantia nigra | 60-69 | 0.774 | 0.461 | 0.803 | 0.628 |
| Brain-Amygdala | 60-69 | 0.736 | 0.480 | 0.760 | 0.626 |
| Brain-Nucleus accumbens basal ganglia | 40-49 | 0.564 | 0.598 | 0.561 | 0.621 |
| Brain-Nucleus accumbens basal ganglia | 60-69 | 0.561 | 0.539 | 0.563 | 0.606 |
| Brain-Hypothalamus | 30-39 | 0.555 | 0.52 | 0.558 | 0.604 |

### 3.2 Comparison between SVM models built by selected and full feature sets

SVM models were built and evaluated using 19 selected and full features (Table 2 and Figure 2). The AUC values for the SVM model based on 19 selected features (0.74 [0.690-0.790]) and full feature sets (0.730 [0.678-0.769]) were very similar. To evaluate model robustness, we also constructed these modes based on the mean expression level of each gene in the tissue type age group. In addition, to examine the potential effects of sex, SVM models were constructed based on the expression data from male and female samples. The results are summarized in Supplementary Table S3. There are no significant differences when mean values were utilized compared to median values. The model performance based on males or females was also very similar to that of models constructed using all samples.

**Table 2.** The average accuracy, sensitivity, specificity and AUC of the two models based on ten-fold cross validation

|  | selected feature set | full feature set |
| --- | --- | --- |
| Accuracy | 0.756±0.016 | 0.754±0.023 |
| Sensitivity | 0.588±0.069 | 0.500±0.054 |
| Specificity | 0.772±0.016 | 0.778±0.021 |
| AUC(95%CI) | 0.740(0.690-0.790) | 0.730(0.678-0.769) |

**Figure 2.**
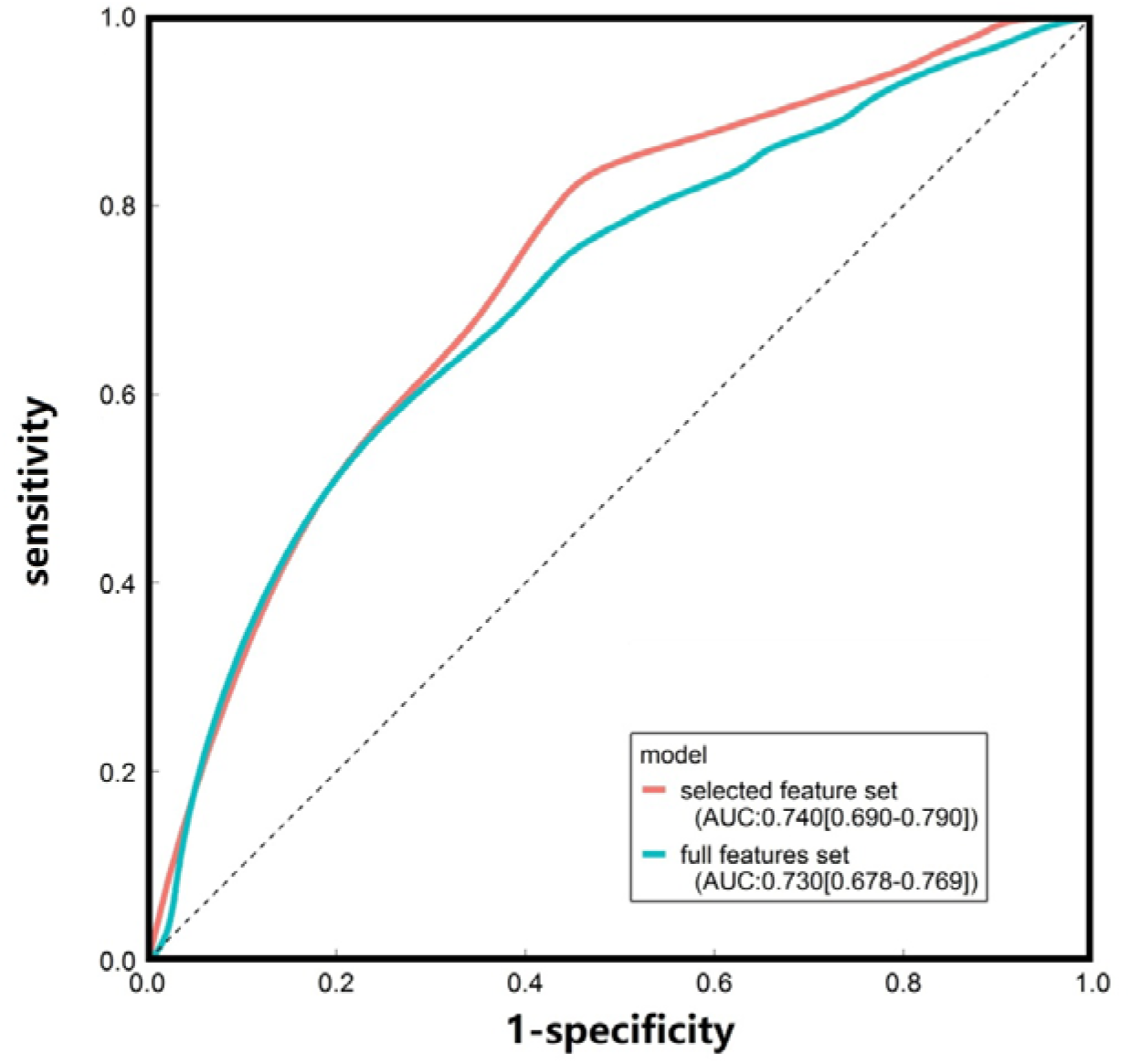
ROC curves of the SVM models constructed based on the median gene expression levels in different tissue-age groups

### 3.3 Risk genes of AD predicted by the SVM model

Based on the SVM models constructed using tissue-age-specific gene expression data, the risk contributions to AD onset and development were evaluated for 10,798 genes that were not included in the model construction and evaluations (the external gene set). Fifteen genes predicted to be risk genes for AD with a probability greater than 90% were obtained (Table 3). *GUCY1B3* had the highest confidence score as a risk gene for AD (0.93). To further investigate this gene set, we examined the gene expression patterns of these 15 genes in the human brain and made a heatmap showing in Supplementary Figure S2. In addition, 191 risk genes for AD with a probability greater than 80% are shown in Supplementary Table S4.

**Table 3.** Genes predicted by the SVM model with their confidence score, location, length(bp) and biotype

| Genes | Confidence | Location | Length (bp) | Type |
| --- | --- | --- | --- | --- |
| *SIX3* | 0.911 | 2p21 | 4,370 | protein coding |
| *EFEMP1* | 0.904 | 2p16.1 | 58,197 | protein coding |
| *GUCY1B3* | 0.939 | 4p32.1 | 48,820 | protein coding |
| *MTPN* | 0.921 | 7q33 | 50,600 | protein coding |
| *ACTR3B* | 0.904 | 7q36.2 | 311,231 | protein coding |
| *BAG3* | 0.932 | 10q26.11 | 26,440 | protein coding |
| *INPP5A* | 0.924 | 10q26.3 | 245,697 | protein coding |
| *LRRC10B* | 0.917 | 11q12.2 | 2,270 | protein coding |
| *ELMOD1* | 0.914 | 11q22.3 | 75,771 | protein coding |
| *DRD2* | 0.900 | 11q23.2 | 66,087 | protein coding |
| *GABRA5* | 0.922 | 15q12 | 82,490 | protein coding |
| *PITPNM3* | 0.916 | 17p13.2-p13.1 | 105,293 | protein coding |
| *SEZ6* | 0.909 | 17q11.2 | 51,540 | protein coding |
| *ICAM5* | 0.922 | 19p13.2 | 7,428 | protein coding |
| *CSDC2* | 0.916 | 22q13.2 | 16,732 | protein coding |

## 4 Discussion

In the present study, we propose a novel machine-learning-based analysis pipeline using data extracted from the GTEx database to prioritize candidate AD risk genes. The performance measured by the AUC of the SVM models was promising, and a list of 15 candidate AD risk genes was presented according to the prediction model. In the last decade, several studies have been published to identify candidate AD risk genes, and most of these studies were based on protein–protein interaction (PPI) networks to identify hub genes using GWA data. The model performance measured by the AUC of these previous studies ranged from 0.63–0.78 depending on different settings. Unlike these previous studies, the STGE framework was used to predict AD candidate genes based on the spatial and temporal features of AD risk gene expression. The performance of our model (AUC=0.74) was comparable to that of previous studies. In this sense, the present study proposed and validated an alternative framework for prioritizing risk genes for AD. In the future, an analysis framework integrating information from gene expression features and PPI network properties might be a promising method to further promote the accuracy and effectiveness of prediction models for prioritizing candidate AD risk genes.

Although most patients with AD experience the first symptom in their mid-60s, previous studies have indicated that changes in the molecular levels occur at a much earlier stage [17-18]. A previously published family-based longitudinal study has shown that familial AD may have a long prodromal phase of several years [19]. A recent cohort study also indicated that plasma phospho-tau181 levels were much higher from 16 years prior to the onset of AD symptoms in AD patients with specific DNA mutations [20-21]. The results of the current study offer new evidence at the gene expression level for prodromal changes in AD patients. Although AD is a late-onset disorder, more than half of the selected features were obtained from sample providers before the age of 60 years. Five of the 19 features, including tissues of the anterior cingulate cortex, putamen basal ganglia, caudate basal ganglia, cerebellum, and hypothalamus, were obtained from providers who are 30–39 years old. In accordance with multiple lines of previous evidence, these findings indicate that molecular-level changes might be identified several years before early symptoms appear in patients with AD. Nevertheless, since a couple of the AD risk genes used in this study were extracted from studies focusing on early-onset AD, we need to be cautious in interpreting these results. Future research using longitudinal data might provide more clues for identifying prodromal biomarkers for AD and, in turn, shed light on early screening and prevention of this complex neurodegenerative disorder.

Among the 15 candidate genes identified through STGE, a few are of particular interest. Sine oculis homeobox homolog 3 (*SIX3*) encodes a type of transcription factor belonging to the sine oculis homeobox transcription factor family [22]. Multiple lines of evidence based on animal models have linked this locus to brain development [22-23]. A recent GWA study associated genetic polymorphisms of *SIX3* with math ability, and its weakening was considered a sign of the progression of AD patients [24]. Actin-related protein 3B (*ACTR3B*) encodes a member of the actin-related protein (ARP) family, which might regulate and induce cell shape changes and motility [25]. Several previous studies have linked *ACTR3B* to brain aging progression, although no direct GWA study has validated the connection between genetic polymorphisms of these loci and AD [25-26]. In addition, multiple animal models and population-based evidence have been published for dopamine receptor D2 (*DRD2*) and gamma-aminobutyric acid type A receptor subunit alpha 5 (*GABRA5*) being associated with brain-related disorders and traits, including schizophrenia, bipolar disorder, Parkinson ‘s disorder, and neurotransmission [27-30]. In a recent study, Blum *et al*. concluded that the *DRD2* Taq1A A1 allele might increase the risk of Alzheimer’s aging in African Americans by integrating and reviewing previously published data [31]. Additionally, the genes BAG Cochaperone 3 (*BAG3*), inositol polyphosphate-5-phosphatase A (*INPP5A*), seizure related 6 homolog (*SEZ6*), and intercellular adhesion molecule 5 (*ICAM5*) are involved in the progression of AD has been proposed in several functional studies using animal models [32-36]. Within these genes, through proteomic study, *BAG3* may affect AD by influencing the interpretation of Aβ and tau protein, and patients with AD have much lower levels of *SEZ6* in their cerebrospinal fluid than those without dementia [37-38]. Further *in vivo* and *in vitro* studies are needed to validate the functional connections between the risk of AD and the genes on the predicted list.

The current study has several limitations. First, there is still much space for the promotion of STGE, although the performance of STGE is comparable to that of previous models based on PPI network properties. In addition, as bioinformatics data mining is based on publicly available databases, the completeness of the current work might be limited owing to data availability. The gene expression data in the brain substantia nigra in the age group of 30–39 years were unavailable from the database; therefore, this feature was not included in the model construction and evaluation. Furthermore, only protein-coding genes were examined in the current study, although non-coding RNA have been shown to play an important role in the pathogenesis of complex disorders [39].

In summary, in the present study, an efficient analysis framework based on spatial and temporal features of gene expression was proposed to prioritize AD risk genes. The newly proposed framework performed comparably to previous prediction methods based on PPI network properties. A list of 15 candidate genes for AD risk was also generated to provide data support for further studies on the genetic etiology of AD.

## Data Availability

The data that support the findings of this study are openly available from the databases described in the Methods part. The codes of this study can be found at https://doi.org/10.5281/zenodo.7553711.

## Acknowledgement

We would thank Mrs. Yingying Wei who has provided insightful suggestions and significantly promoted the manuscript.

