## Supplementary Table S1 for "Prioritization of risk genes for Alzheimer’s disease: an analysis framework using spatial and temporal gene expression data in the human brain based on support vector machine"

**Supplementary Table S1.** The number of samples in each tissue-age group.

| Types of Brain Tissues | Age | | | | |
| --- | --- | --- | --- | --- | --- |
|  | 20-29 | 30-39 | 40-49 | 50-59 | 60-69 |
| Cerebellum | 7 | 3 | 13 | 49 | 54 |
| Cortex | 5 | 3 | 16 | 43 | 46 |
| Anterior cingulate cortex | 3 | 1 | 11 | 27 | 37 |
| Hippocampus | 5 | 2 | 11 | 27 | 34 |
| Substantia nigra | 4 | 0 | 7 | 20 | 25 |
| Caudate (basal ganglia) | 5 | 2 | 14 | 39 | 48 |
| Cerebellar Hemisphere | 7 | 3 | 10 | 34 | 42 |
| Frontal Cortex | 3 | 3 | 7 | 34 | 37 |
| Hypothalamus | 4 | 3 | 7 | 31 | 32 |
| Nucleus accumbens (basal ganglia) | 6 | 2 | 12 | 36 | 42 |
| Putamen (basal ganglia) | 5 | 1 | 9 | 37 | 39 |
| Spinal cord | 4 | 1 | 5 | 25 | 24 |
| Amygdala | 4 | 2 | 6 | 25 | 25 |
