## Supplementary Table S2 for "Prioritization of risk genes for Alzheimer’s disease: an analysis framework using spatial and temporal gene expression data in the human brain based on support vector machine"

**Supplementary Table S2.** Information of the 64 features for brain related tissues.

| Human Tissues | Age |
| --- | --- |
| Brain - Amygdala | 20-29 |
| Brain - Amygdala | 30-39 |
| Brain - Amygdala | 40-49 |
| Brain - Amygdala | 50-59 |
| Brain - Amygdala | 60-69 |
| Brain - Anterior cingulate cortex (BA24) | 20-29 |
| Brain - Anterior cingulate cortex (BA24) | 30-39 |
| Brain - Anterior cingulate cortex (BA24) | 40-49 |
| Brain - Anterior cingulate cortex (BA24) | 50-59 |
| Brain - Anterior cingulate cortex (BA24) | 60-69 |
| Brain - Caudate (basal ganglia) | 20-29 |
| Brain - Caudate (basal ganglia) | 30-39 |
| Brain - Caudate (basal ganglia) | 40-49 |
| Brain - Caudate (basal ganglia) | 50-59 |
| Brain - Caudate (basal ganglia) | 60-69 |
| Brain - Cerebellar Hemisphere | 20-29 |
| Brain - Cerebellar Hemisphere | 30-39 |
| Brain - Cerebellar Hemisphere | 40-49 |
| Brain - Cerebellar Hemisphere | 50-59 |
| Brain - Cerebellar Hemisphere | 60-69 |
| Brain - Cerebellum | 20-29 |
| Brain - Cerebellum | 30-39 |
| Brain - Cerebellum | 40-49 |
| Brain - Cerebellum | 50-59 |
| Brain - Cerebellum | 60-69 |
| Brain - Cortex | 20-29 |
| Brain - Cortex | 30-39 |
| Brain - Cortex | 40-49 |
| Brain - Cortex | 50-59 |
| Brain - Cortex | 60-69 |
| Brain - Frontal Cortex (BA9) | 20-29 |
| Brain - Frontal Cortex (BA9) | 30-39 |
| Brain - Frontal Cortex (BA9) | 40-49 |
| Brain - Frontal Cortex (BA9) | 50-59 |
| Brain - Frontal Cortex (BA9) | 60-69 |
| Brain - Hippocampus | 20-29 |
| Brain - Hippocampus | 30-39 |
| Brain - Hippocampus | 40-49 |
| Brain - Hippocampus | 50-59 |
| Brain - Hippocampus | 60-69 |
| Brain - Hypothalamus | 20-29 |
| Brain - Hypothalamus | 30-39 |
| Brain - Hypothalamus | 40-49 |
| Brain - Hypothalamus | 50-59 |
| Brain - Hypothalamus | 60-69 |
| Brain - Nucleus accumbens (basal ganglia) | 20-29 |
| Brain - Nucleus accumbens (basal ganglia) | 30-39 |
| Brain - Nucleus accumbens (basal ganglia) | 40-49 |
| Brain - Nucleus accumbens (basal ganglia) | 50-59 |
| Brain - Nucleus accumbens (basal ganglia) | 60-69 |
| Brain - Putamen (basal ganglia) | 20-29 |
| Brain - Putamen (basal ganglia) | 30-39 |
| Brain - Putamen (basal ganglia) | 40-49 |
| Brain - Putamen (basal ganglia) | 50-59 |
| Brain - Putamen (basal ganglia) | 60-69 |
| Brain - Spinal cord (cervical c-1) | 20-29 |
| Brain - Spinal cord (cervical c-1) | 30-39 |
| Brain - Spinal cord (cervical c-1) | 40-49 |
| Brain - Spinal cord (cervical c-1) | 50-59 |
| Brain - Spinal cord (cervical c-1) | 60-69 |
| Brain - Substantia nigra | 20-29 |
| Brain - Substantia nigra | 40-49 |
| Brain - Substantia nigra | 50-59 |
| Brain - Substantia nigra | 60-69 |
