## Supplementary Table S3 for "Prioritization of risk genes for Alzheimer’s disease: an analysis framework using spatial and temporal gene expression data in the human brain based on support vector machine"

**Supplementary Table S3.** Model constructions and evaluations based on both median and mean values and gender stratifications.

| Classes of Models | Accuracy | Sensitivity | Specificity | AUC(95%CI) |
| --- | --- | --- | --- | --- |
| Selected Feature-Median | 0.756 | 0.588 | 0.772 | 0.740(0.690-0.790) |
| Full Feature-Median | 0.754 | 0.500 | 0.778 | 0.730(0.678-0.769) |
| Selected Feature-Mean | 0.742 | 0.529 | 0.762 | 0.718(0.668-0.769) |
| Full Feature-Mean | 0.738 | 0.539 | 0.757 | 0.716(0.669-0.762) |
| Selected Feature-Males | 0.726 | 0.578 | 0.741 | 0.728(0.682-0.774) |
| Selected Feature -Females | 0.762 | 0.490 | 0.788 | 0.734(0.691-0.778) |
