## Supplementary Table S4 for "Prioritization of risk genes for Alzheimer’s disease: an analysis framework using spatial and temporal gene expression data in the human brain based on support vector machine"

**Supplementary Table S4.** 191Risk genes of AD with a probability greater than 80%.

| Names of genes | Probability |
| --- | --- |
| *GUCY1B3* | 0.939 |
| *BAG3* | 0.932 |
| *INPP5A* | 0.924 |
| *GABRA5* | 0.922 |
| *ICAM5* | 0.922 |
| *MTPN* | 0.921 |
| *LRRC10B* | 0.917 |
| *CSDC2* | 0.916 |
| *PITPNM3* | 0.916 |
| *ELMOD1* | 0.914 |
| *SIX3* | 0.911 |
| *SEZ6* | 0.909 |
| *EFEMP1* | 0.904 |
| *ACTR3B* | 0.904 |
| *DRD2* | 0.900 |
| *ANP32E* | 0.899 |
| *HMGN1* | 0.897 |
| *RBP4* | 0.895 |
| *MAPK1* | 0.892 |
| *ADORA2A* | 0.892 |
| *AKAP12* | 0.891 |
| *EDNRB* | 0.891 |
| *CAP2* | 0.890 |
| *PKIA* | 0.889 |
| *OXCT1* | 0.886 |
| *SPATA20* | 0.884 |
| *RBM25* | 0.883 |
| *GAD2* | 0.882 |
| *LHFP* | 0.881 |
| *RIMS4* | 0.880 |
| *PSD3* | 0.879 |
| *LRP1* | 0.878 |
| *CDC25B* | 0.876 |
| *ULK1* | 0.876 |
| *NRSN1* | 0.874 |
| *CACNG3* | 0.874 |
| *TRIM9* | 0.873 |
| *POGZ* | 0.872 |
| *SPEF1* | 0.869 |
| *GOLGA8B* | 0.869 |
| *GSTM1* | 0.868 |
| *VARS2* | 0.865 |
| *ARHGEF12* | 0.863 |
| *CHRM1* | 0.863 |
| *WAC* | 0.862 |
| *C2orf80* | 0.862 |
| *RBM5* | 0.861 |
| *SNCA* | 0.860 |
| *PPAP2B* | 0.860 |
| *UNC13A* | 0.858 |
| *GABRG2* | 0.858 |
| *CIC* | 0.858 |
| *SLC4A3* | 0.857 |
| *PRRC2B* | 0.855 |
| *G3BP2* | 0.855 |
| *UPF3A* | 0.854 |
| *MPRIP* | 0.852 |
| *LSAMP* | 0.850 |
| *MEF2D* | 0.850 |
| *EIF3E* | 0.850 |
| *CA12* | 0.850 |
| *TTYH3* | 0.850 |
| *ANO3* | 0.849 |
| *SPON1* | 0.849 |
| *CALB1* | 0.848 |
| *SUN1* | 0.848 |
| *ADRA2C* | 0.846 |
| *CRY2* | 0.846 |
| *CELF4* | 0.846 |
| *GSTA4* | 0.845 |
| *STAT5B* | 0.844 |
| *PARM1* | 0.844 |
| *SCN2B* | 0.843 |
| *LPPR3* | 0.842 |
| *LPHN1* | 0.841 |
| *RNF146* | 0.841 |
| *LZTS1* | 0.840 |
| *WBSCR17* | 0.839 |
| *CACNG4* | 0.838 |
| *RGS9* | 0.838 |
| *R3HDM2* | 0.838 |
| *SLC1A4* | 0.838 |
| *KIAA1211L* | 0.837 |
| *UBE2G2* | 0.837 |
| *FAM171B* | 0.837 |
| *RBM6* | 0.836 |
| *MBTPS1* | 0.836 |
| *PRKAR2B* | 0.835 |
| *SLC32A1* | 0.835 |
| *RAF1* | 0.835 |
| *FAM219B* | 0.835 |
| *CLASP2* | 0.835 |
| *SALL2* | 0.835 |
| *HP1BP3* | 0.835 |
| *TSC2* | 0.834 |
| *ILF3* | 0.833 |
| *MYO5A* | 0.833 |
| *RAP1GAP2* | 0.832 |
| *ARHGEF17* | 0.832 |
| *DRD1* | 0.831 |
| *EIF2A* | 0.830 |
| *ARHGEF9* | 0.830 |
| *ST8SIA3* | 0.829 |
| *PLCG1* | 0.829 |
| *RAPGEF1* | 0.829 |
| *GABBR2* | 0.827 |
| *ARRB1* | 0.827 |
| *POLR2A* | 0.826 |
| *EXOSC10* | 0.826 |
| *MAP2K1* | 0.825 |
| *PRPF8* | 0.824 |
| *ITPR1* | 0.824 |
| *SAMD14* | 0.824 |
| *ATXN2L* | 0.823 |
| *DLGAP4* | 0.823 |
| *DPP6* | 0.822 |
| *KCNQ2* | 0.822 |
| *SYT13* | 0.822 |
| *CPNE1* | 0.822 |
| *KNDC1* | 0.822 |
| *ATP2B1* | 0.821 |
| *SRRM1* | 0.821 |
| *PJA1* | 0.821 |
| *RNF114* | 0.821 |
| *ABLIM2* | 0.820 |
| *GNAL* | 0.820 |
| *LGI1* | 0.820 |
| *PNMT* | 0.819 |
| *SCN3B* | 0.819 |
| *C16orf58* | 0.818 |
| *SRSF3* | 0.817 |
| *NTSR2* | 0.816 |
| *FRRS1L* | 0.816 |
| *NPEPPS* | 0.816 |
| *MXRA7* | 0.816 |
| *MCFD2* | 0.816 |
| *MGAT3* | 0.816 |
| *PAM* | 0.816 |
| *GRIPAP1* | 0.815 |
| *ANGPTL4* | 0.815 |
| *SUB1* | 0.814 |
| *CD200* | 0.814 |
| *ATF6B* | 0.814 |
| *TESC* | 0.814 |
| *MIAT* | 0.814 |
| *CHD4* | 0.814 |
| *DNM1L* | 0.813 |
| *ADAM23* | 0.813 |
| *MLLT1* | 0.813 |
| *FGF13* | 0.813 |
| *PTPRZ1* | 0.813 |
| *C7orf55-LUC7L2* | 0.813 |
| *ABCB6* | 0.813 |
| *TGOLN2* | 0.813 |
| *SF3B1* | 0.813 |
| *SMARCA2* | 0.812 |
| *ADCY5* | 0.812 |
| *DNAJC6* | 0.812 |
| *LRPPRC* | 0.811 |
| *TMEM47* | 0.811 |
| *LONRF2* | 0.811 |
| *HNRNPH1* | 0.811 |
| *SPRY2* | 0.810 |
| *PRKCB* | 0.810 |
| *CPSF3L* | 0.809 |
| *LDB1* | 0.809 |
| *LMO3* | 0.809 |
| *RPS6KA4* | 0.808 |
| *RABL6* | 0.808 |
| *GLS* | 0.807 |
| *RAB3C* | 0.807 |
| *ACBD7* | 0.806 |
| *FLCN* | 0.806 |
| *ANK2* | 0.806 |
| *GGT7* | 0.805 |
| *ACTN1* | 0.805 |
| *DDX3X* | 0.804 |
| *DCAF8* | 0.804 |
| *TOP2B* | 0.804 |
| *AP2A2* | 0.804 |
| *EDC4* | 0.803 |
| *AMPD2* | 0.803 |
| *PPP6R2* | 0.803 |
| *SPSB3* | 0.803 |
| *F3* | 0.802 |
| *DLST* | 0.802 |
| *SYNGAP1* | 0.802 |
| *CELF2* | 0.801 |
| *CRTC1* | 0.801 |
| *COX7A1* | 0.801 |
| *ID4* | 0.800 |
