## Supplementary figures and images for "Prioritization of risk genes for Alzheimer’s disease: an analysis framework using spatial and temporal gene expression data in the human brain based on support vector machine"

### Supplemental Figure S1

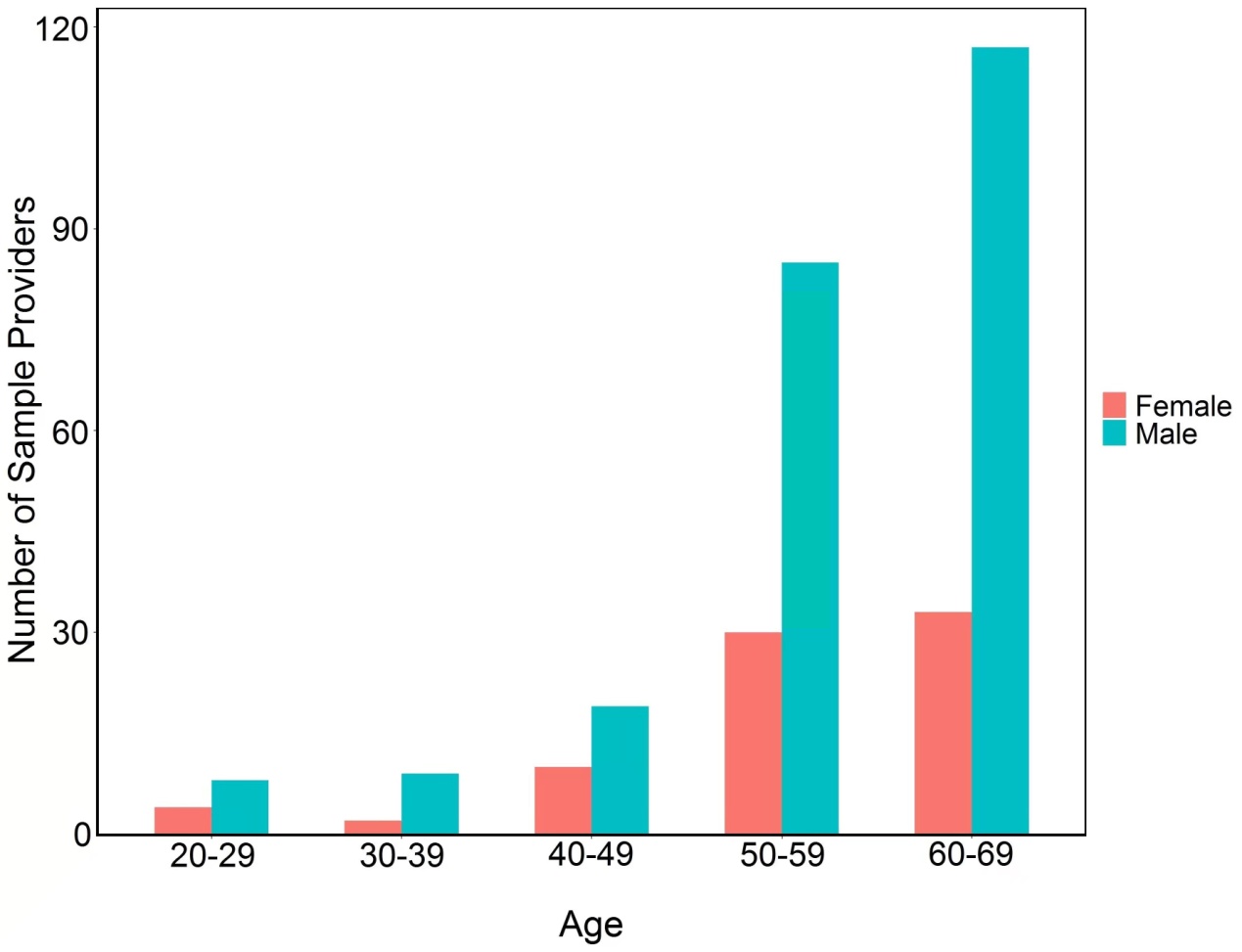

**Supplemental Figure S1.** Sex and age distribution of the tissue sample providers.
