## Supplemental Figure S2 for "Prioritization of risk genes for Alzheimer’s disease: an analysis framework using spatial and temporal gene expression data in the human brain based on support vector machine"

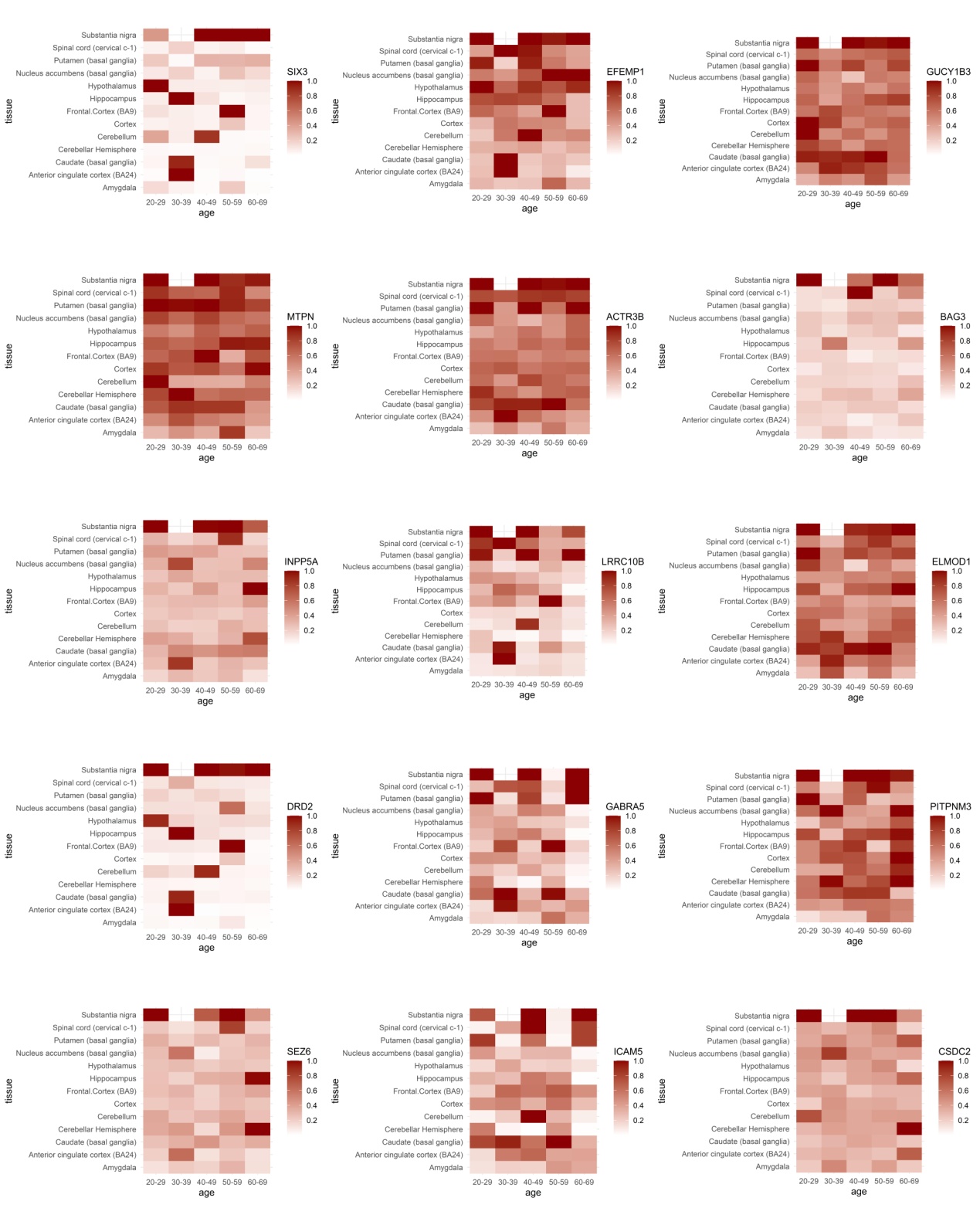

**Supplementary Figure S2.** Heatmap of the gene expression patterns for 15 AD risk genes identified through the SVM models.
